# Aggregating probabilistic predictions of the safety, efficacy, and timing of a COVID-19 vaccine

**DOI:** 10.1101/2021.06.03.21258240

**Authors:** Thomas McAndrew, Juan Cambeiro, Tamay Besiroglu

## Abstract

Safe, efficacious vaccines were developed to reduce the transmission of SARS-CoV-2 during the COVID-19 pandemic. But in the middle of 2020, vaccine effectiveness, safety, and the timeline for when a vaccine would be approved and distributed to the public was uncertain. To support public health decision making, we solicited trained forecasters and experts in vaccinology and infectious disease to provide monthly probabilistic predictions from July to September of 2020 of the efficacy, safety, timing, and delivery of a COVID-19 vaccine. We found, that despite sparse historical data, a consensus—a combination of human judgment probabilistic predictions—can quantify the uncertainty in clinical significance and timing of a potential vaccine. The consensus underestimated how fast a therapy would show a survival benefit and the high efficacy of approved COVID-19 vaccines. However, the consensus did make an accurate prediction for when a vaccine would be approved by the FDA. Compared to individual forecasters, the consensus was consistently above the 50^th^ percentile of the most accurate forecasts. A consensus is a fast and versatile method to build probabilistic predictions of a developing vaccine that is robust to poor individual predictions. Though experts and trained forecasters did underestimate the speed of development and the high efficacy of a SARS-CoV-2 vaccine, consensus predictions can improve situational awareness for public health officials and for the public make clearer the risks, rewards, and timing of a vaccine.

## I. INTRODUCTION

SARS-CoV-2—the infectious agent that causes COVID-19—spread rapidly throughout the world and is responsible for millions of deaths [1–5]. The pandemic has negatively impacted societal, psychological, and economic factors, made those with comorbidities more susceptible to disease, and may have increased health inequities [6–15].

In response, public health officials have offered guidance on non-pharmaceutical interventions (NPI), and have tracked and forecasted the transmission of SARS-CoV-2. However both tracking and NPI efforts have had mixed results [16–23]. Immunization has been shown to be an effective and long term solution to reducing the burden of many different diseases [24, 25]. Computational models predict widespread delivery of effective vaccines in addition to continued adherence to non-pharmaceutical interventions have the potential to reduce the impact of SARS-CoV-2 and COVID-19 [26–28].

Three vaccines to protect against COVID-19 disease have been authorized on an emergency basis by the Food and Drug Administration (FDA): a vaccine made by Pfizer and BioNTech with a reported efficacy of 95%, a vaccine produced by Moderna with an efficacy of 94%, and a single-dose vaccine produced by Johnson and Johnson with an efficacy of 66% [29–31]. Similar to tracking the transmission of disease, the U.S. Centers for Disease Control and Prevention (CDC) tracks the number of allocated, delivered, and administered COVID-19 vaccines in the U.S. to improve the situational awareness of public health decision-makers [32].

But in the middle of 2020 there was no approved COVID-19 vaccine. Vaccines were still in progress and whether a vaccine would be authorized, how safe and effective a vaccine would be, and how fast a vaccine could be produced and delivered to the public was uncertain.

Our goal was to aggregate probabilistic predictions of the safety, efficacy, and timelines of a COVID-19 vaccine from trained, generalist forecasters and experts in vaccinology and infectious disease into a single *consensus* prediction and produce reports to support public health decision-making. We used an online human judgment forecasting platform to collect probabilistic predictions from June 2020 to September 2020 [33].

Computational ensembles during the COVID-19 pandemic have been used to guide the selection and continued evaluation of trial sites for vaccine efficacy studies [34] and have been used to predict the number of individuals fully vaccinated [35]. Human judgment has been applied to predict both epidemiological targets of COVID-19 [36] and aspects of the various vaccination efforts against COVID-19 [37, 38].

A multi-model ensemble combines predictive distributions from individual models to produce a single predictive distribution. Because an ensemble can combine models trained on different sets of data and with different underlying assumptions, an ensemble has the potential to produce a more accurate, better calibrated predictive distribution compared to the individual models that contributed to the ensemble [39, 40]

Aggregating predictions from subject-matter experts and generalist forecasters offers an alternative to computational modeling and can be useful when historical data is sparse, predictions are needed for a small number of targets, and when information needed by public health officials can change rapidly [36, 39, 41, 42]. Human judgment has been applied to generate probabilistic forecasts in several different domains [41–45]. In the field of infectious disease, human judgment has predicted rates of US influenza-like illness [46], West-Nile virus [47] and Malaria [48], and epidemiological targets of the early US trajectory of COVID-19 [36].

Forecasts of vaccine characteristics have the potential to impact public, public health official, and vaccine research and development decision making under uncertainty. Efficacy and safety of vaccine may determine whether an individual volunteers to be inoculated [49–52]. For public health officials, predictions of the time to approval and time to manufacture a vaccine serve as valuable input in supply chain management, including logistics planning, inventory management, material requirements planning [53–55]. There is also a need from those in research and development for probabilistic predictions to help determine which vaccine platforms and pathways should be pursued before others [56–59].

To the best of our knowledge, this work is the first to generate ensemble probabilistic predictions from expert and generalist forecasters on COVID-19 vaccine development and share these with the public and public health decision-makers from June 2020 through September 2020, before the first approved COVID-19 vaccine.

## II. METHODS

### II.1. Survey timeline

Subject matter experts (SMEs) and trained forecasters (definition below) participated in four surveys from June 15th, 2020 to August 30th, 2020. SMEs and forecasters were asked to predict aspects of safety, efficacy, and delivery of a COVID-19 vaccine (see [33] to view all four summary reports, questions asked of forecasters and collected prediction data used in this work).

Subject matter experts and trained forecasters were solicited by sending personal emails (see Supp. S1 for a template email sent to subject matter experts). Solicitation started on June 3, 2020 and ended on July 8, 2020.

The first two weeks of each month (the 1st to the 14th) was used to develop questions that could address changing information about a COVID-19 vaccine.

Forecasters received a set of questions on the 15th of each month and from the 15th to the 25th could submit predictions using the Metaculus platform [60]. Forecasters made a first prediction and, as many times as they wished, could revise their original prediction between the 15th and 25th. To reduce anchoring bias [61], between the 15th and the 20th forecasters made predictions without knowledge of other forecaster’s predictive densities. From the 20th to the 25th a consensus predictive density—an equally weighted combination of predictive densities from subject matter experts and trained forecasters—was available to forecasters.

### II.2. Forecasters

We defined a subject matter expert as someone with training in the fields of molecular and cellular biology, microbiology, virology, biochemistry, and infectious diseases, and who has several years of experience in vaccine, antiviral, and/or biological research related to infectious agents and kept up-to-date with vaccine and antiviral research specifically focused on SARS-CoV-2/ COVID-19. Subject matter experts were trained in biological sciences but were not required to have had prior experience making accurate, calibrated probabilistic predictions.

We defined a trained forecaster as someone who ranked in the top 1% out all Metaculus forecasters (approximately 15,000 forecasters), according to a Metaculus point system, and who has made consistent predictions on the Metaculus forecasting platform for a minimum of one year. Past work has shown that individuals who make consistent and accurate forecasts of one set of targets may be able to apply their forecasting skills to other targets [62–64]. Trained forecasters were not required to have a background in vaccines, biology, or infectious diseases, but they were required to have a history of making accurate predictions across a variety of topics and forecasting tournaments.

### II.3. Vaccine-related questions

Forecasters were asked questions that fell into four categories: safety, efficacy, timing and delivery, and urgent matters (see Suppl. Table S1 for a list of all questions asked over all four surveys).

All questions asked of forecasters contained background information related to the target of interest, the question itself, and a detailed paragraph that described how we will determine the true, final value used to score predictive distributions.

Safety questions asked about survival rates related to vaccination. In particular, forecasters were asked to predict the probability that more than 10 patients experience a serious adverse event from a COVID-19 vaccine within one year of the date that vaccine was approved, and when a randomized controlled trial that tests a COVID-19 vaccine compared to a suitable control will show a statistically significant survival benefit.

Efficacy questions aimed to estimate the relative difference in attack rate between those vaccinated and those unvaccinated for the (at the time) ongoing trial to test the ChAdOx1 vaccine, for an approved vaccine under standard regulatory guidance or an emergency use authorization, and for a vaccine using one of several different viral platforms.

Timing and delivery questions asked forecasters to predict when the first SARS-CoV-2 vaccine would be approved in the United States or in the European Union, and to compare differences between the date that the FDA would approve a SARS-CoV-2 vaccine under a standard regulatory process versus expedited (emergency) process. Forecasters were also asked to predict the elapsed time between when a vaccine would be approved and when 100M doses would be manufactured.

In addition to safety, efficacy, and timing and delivery, forecasters were asked to predict the number of vaccine candidates in both clinical and pre-clinical development.

Questions were designed to communicate the risks (efficacy and safety) of being inoculated with a COVID-19 vaccine candidate; provide public officials a potential timeline of vaccine approval so that they can prepare; present objective, falsifiable predictions of vaccine development from an expert crowd; and decrease potential misinformation available to the public.

### II.4. Forecasting platform

Forecasters submitted predictive densities by accessing the Metaculus platform (Metaculus), an online forecasting platform that allows users to submit predictive densities and comments related to a proposed question. Metaculus stores individual predictions and comments for each question and when the ground truth for a question is available the platform scores individual predictions and keeps a history of each forecaster’s average score across all questions for which they have submitted predictions. The Metaculus platform allows participants to visualize their proposed predictive density both as probability density and cumulative density functions.

A private subdomain was created on Metaculus that allowed only subject matter experts and select trained forecasters to submit predictions and comments on these COVID-19 vaccine questions. Forecasters were encouraged, but not required, to answer all questions. When a user accesses Metaculus they are presented with a list of questions for which they can submit probabilistic predictions. For each question, the forecaster was presented background information about the specific question, including resources judged by the authors to be relevant and informative. Each question also contained a detailed statement of the resolution criteria—the criteria that describes, as precisely as possible, how the ground-truth would be determined.

Below the question, forecasters are presented with a tool to form a predictive density as a weighted mixture of up to five logistic distributions (See Fig. 1). The logistic distribution resembles a normal distribution but has heavier tails.

**FIG. 1:**
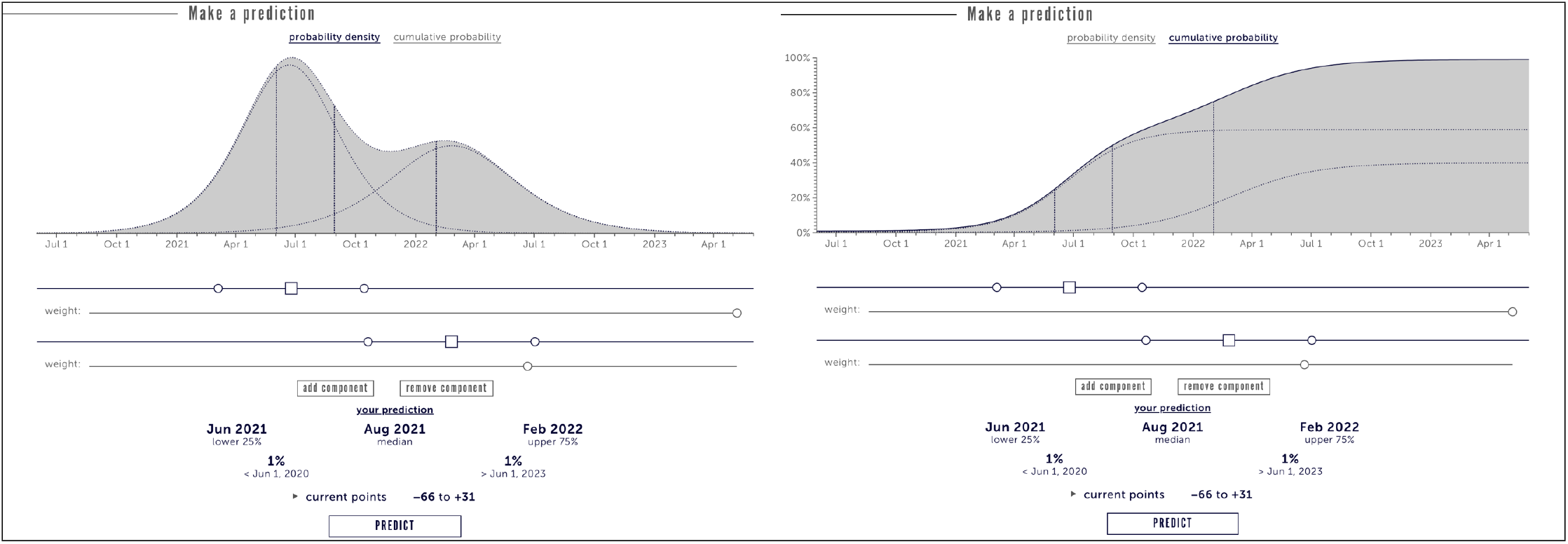
The Metaculus prediction interface which records forecaster’s probabilistic predictions. (Left) The probability density view (Right) The cumulative density view. The prediction interface allows users to shift and scale up to five logistic distributions (in this example two distributions are used) and increase or decrease the weight given to each distribution. The median, 25^th^, and 75^th^ percentiles are displayed to the forecaster.

### II.5. Individual predictions

Experts and trained forecasters submit a probabilistic density as a convex combination of up to five logistic distributions. Specifically, the ensemble probabilistic prediction *f*_*m*_ for the *m*^th^ forecaster is given by:

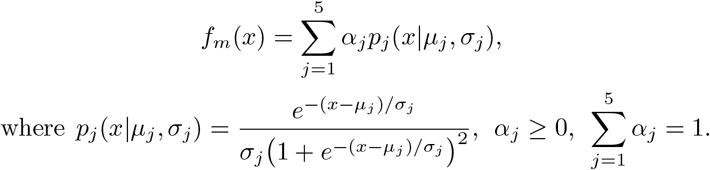

When generating a prediction, participants specify at minimum one logistic distribution by moving a slider that corresponds to *µ* and compressing or expanding the same slider that corresponds to *σ*. If a participant decides to add a second (or 3^rd^, 4^th^, and 5^th^) logistic distribution they can click “add a component” for a second slider allowing the participant to “shift and scale” this additional logistic distribution. Two “weight” sliders also appear under each “shift and scale” slider that allows the participant to control the weights (*α*_1_, *α*_2_,…, *α*_5_) of each individual logistic distribution.

Forecasters can assign probabilities over a domain of pre-specified possible outcomes. The domain (*D*) is typically a closed interval of the real number line where the lower (*L*) and upper bounds (*U*) of the interval are chosen to contain all possible outcomes (*D* = [*L, U*]). In some cases an additional outcome allows forecasters to assign probability to an open-ended outcome falling outside the upper or lower bound of the domain (*D* = *𝓁*∪ [*L, U*] ∪*υ*). For example, for the question “When will a COVID-19 vaccine show an efficacy of greater than or equal to 70%?” a forecaster could assign a probability density from Aug, 2020 to April, 2023 and to the outcome “later than April, 2023”.

A submitted individual forecaster ensemble probabilistic prediction is stored by the forecasting platform as an array of 200 density values evaluated at 200 equally spaced points from the minimum to maximum allowable values (i.e. 200 equally spaced values in [*L, U*]). For questions with no upper or lower bound, densities are still stored over the interval [*L, U*] with the understanding that because the interval [*L, U*] does not cover all possible events the probability over [*L, U*] is less than one.

### II.6. Consensus building

A consensus predictive density is a convex combination (often called a linear pool [65]) of probabilistic predictions from individuals

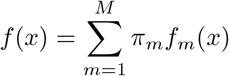

where *M* is the number of forecasters who contributed a predictive distribution and *f*_*m*_ is the *m*^th^ forecaster’s predictive density with an associated consensus weight *π*_*m*_. The sum of weights for all *M* forecasters must sum to one 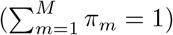. For all consensus forecasts we chose to assign equal weights to each forecaster (i.e. 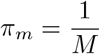),as we had little a-priori reason to assign differential weights to participants.

### II.7. Scoring

We chose to score forecasts using the logarithmic (log) score [66, 67]. The log score assigns the logarithm of the density value corresponding to the eventual true value (*t*) of a target of interest.

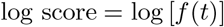

where *f* is the predictive density submitted by an individual forecaster or consensus. Log scores take values from negative to positive infinity. The worst possible log score a forecaster can receive is negative infinity (earned when the density assigned to the actual outcome value is zero), and the best possible score is positive infinity (earned when the density assigned to the actual outcome approaches positive infinity).

The log score is a proper scoring rule. A proper scoring rule is optimized when a forecaster submits the true density over potential values of a target, disincentivizing a forecaster from submitting a predictive density that does not accurately represent their true uncertainty over potential outcomes [68, 69].

Scaled ranks were also reported for logscores. Given a set of *N* log scores, the scaled rank assigns a value of 1*/N* to the smallest log score, a value of 2*/N* to the second smallest log score, and so on, assigning a value of 1 to the highest log score.

### II.8. Statistical inference and testing

A mixed effects regression model was fit to log scores from experts, trained forecasters, and consensus models (considered a forecasters) for all questions with ground truth data. The model is

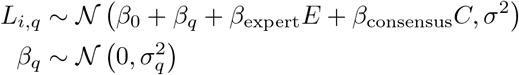

where *L*_*iq*_ is the log score generated by forecaster *i* who made a prediction for question *q, E* is a binary variable that identifies whether a forecaster was an expert (*E* = 1) or not (*E* = 0), *C* is a binary variable that indicates whether a forecaster was a consensus model (*C* = 1) or not (*C* = 0). and *β*_*q*_ is a normally distributed, random intercept, with standard deviation *σ*_*q*_ that accounts for the tendency for log scores to be clustered within each question.

All statistical hypothesis tests are two-sided and a pvalue less than 0.05 is considered statistically significant.

### II.9. Comments and information exchange

Before, during, and after a question is available for forecasting, participants can post comments. Comments are used to clarify question text, for a forecaster to explain their rationale behind a forecast, and for signaling other potential forecasters about a potentially important data source (forecasters can notify one another by adding an “@” to a specific forecaster’s username).

Forecasters can also interact with one another through a consensus distribution generated by Metaculus which was revealed for each question on the 25th of each month, approximately half way through the total time left to make predictions. The consensus distribution was updated after the 25th as individuals made new predictions or if an individual revised their previous prediction.

## III. RESULTS

### III.1. Participation and response rate of the crowd

Four monthly surveys were conducted in June, July, August, and September of 2020. A total of 10 individual experts and 11 trained forecasters participated in at least one of the four surveys. On average 6 experts participated (made at least one prediction for one question) per survey and an average of 8 trained forecasters participated per survey. The median number of hours after the survey was open until the first prediction by an expert was 12 hours and 9 hours for trained forecasters, and the last prediction by an expert was on average 6 hours and for a trained forecaster 2 hours before survey close.

Experts and trained forecasters (the crowd) were asked on average 6.5 questions and made on average 132.5 unique and revised predictions per survey. In June we asked 6 questions and received 154 total (unique and revised) predictions (26 predictions per question), in July we asked 7 questions and received 148 predictions (21 predictions per question), in August we asked 8 questions and received 153 predictions (19 predictions per question), and in September we asked 5 questions and received 75 predictions (15 predictions per question).

Comments were made on 21 out of 26 questions (80.7%) with an average of 2.2 comments per question across all four surveys and a maximum number of comments of 8 on the question asked in June “When will a SARS-CoV-2 vaccine candidate demonstrate ≥ 70% efficacy?”.

### III.2. Efficacy

The consensus median prediction for when a vaccine would show an efficacy above 70% was Aug. 13, 2021 (IQR: [Mar. 2021, Feb., 2022]) and the consensus assigned a 0.12 probability to a COVID-19 vaccine reporting an efficacy of 70% or higher between June 30th, 2020 and Dec. 10, 2020. The consensus median prediction for the efficacy of the ChAdOx1 vaccine was 55.2% (80CI: [7.5%, 83.6%]), the consensus expected a higher reported efficacy for a vaccine that underwent a standard regulatory process (median: 67%; IQR: [58%, 75%]) compared to an expedited emergency use authorization (median: 50%; IQR: [37%, 63%]), and the consensus assigned the highest median efficacy to a vaccine using a protein subunit platform (median: 69%; IQR: [54%, 79%]) compared to a non-replicating viral platform, inactiviated virus platform, and DNA/RNA platform. We asked trained forecasters and experts eight questions, two in June, four in July, and two in August, related to the efficacy of a COVID-19 vaccine (Fig. 2) and received 160 predictions (20 predictions per question on average).

**FIG. 2:**
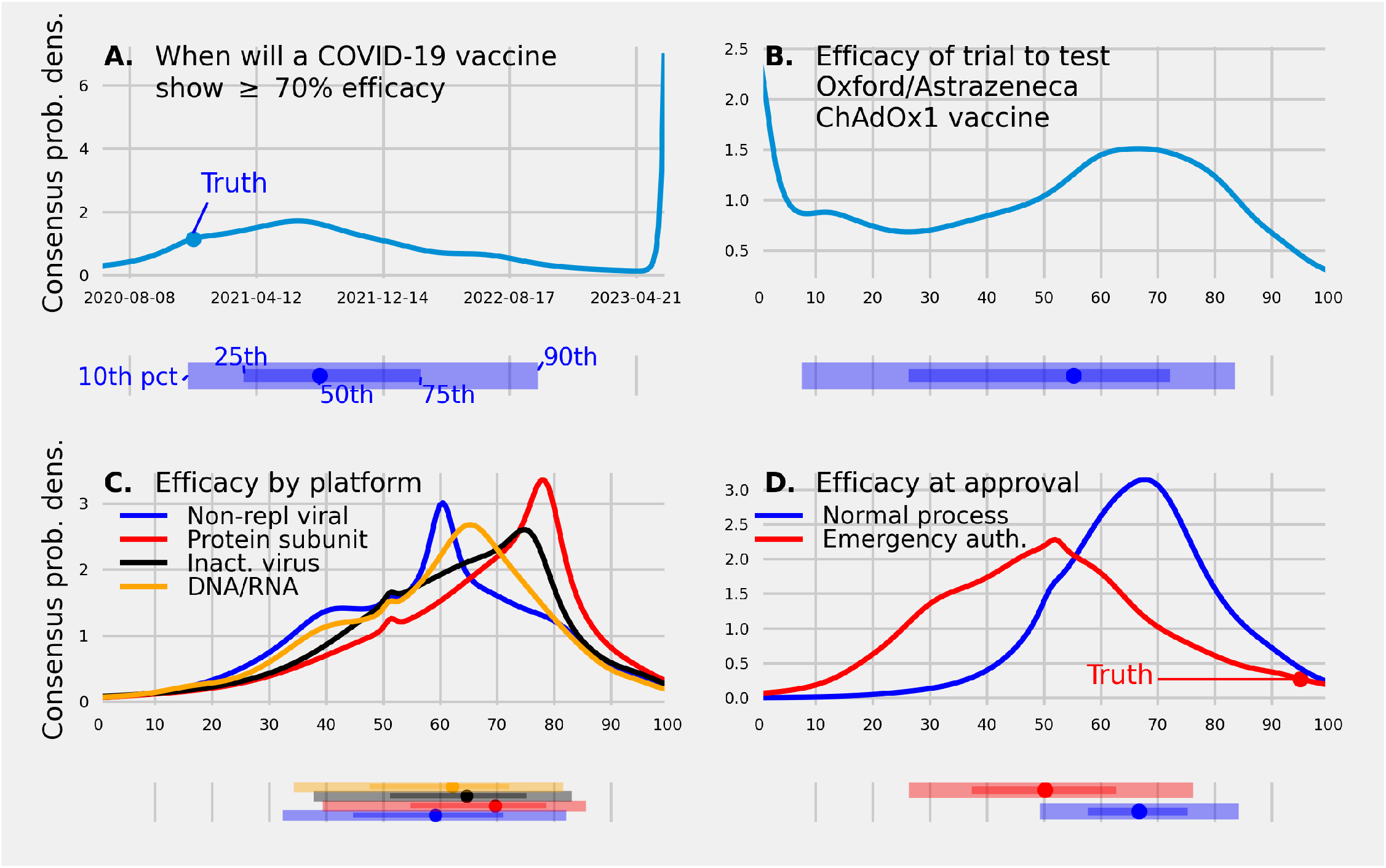
(A.) A consensus predictive density made in June, 2020 of the date when a COVID-19 vaccine will demonstrate an efficacy of 70% or greater. The consensus assigned a 0.12 probability to a vaccine showing a 70% or greater efficacy by Dec. 10, 2020, the date the Pfizer and BioNTech vaccine was approved. (B.) A consensus predictive density made in June, 2020 of the efficacy reported from the trial testing the ChAdOx1 vaccine (C.) A consensus predictive density made in July, 2020, of the efficacy of a vaccine based on four different platforms (D.) A consensus predictive density made in August, 2020 of the efficacy of a vaccine at approval under a standard regulatory process and emergency use authorization. Under each predictive density is the corresponding 10th, 25th, 50th (median), 75th, and 90th quantile. The true values, if available, are represented as a filled circle. A consensus of experts and trained forecasters made probabilistic predictions that compared vaccine efficacy between different regulatory mechanisms and between different vaccine delivery methods, gave a time-frame for when an efficacious vaccine will be approved, and made a testable prediction of the efficacy of a specific trial of interest.

The consensus median prediction made in June, 2020 for when a vaccine would show an efficacy above 70% was Aug. 13, 2021, the mode was July 1, 2021, and the 90% confidence interval (CI) was [Oct., 2020, May, 2023] (Fig. 2A.). The consensus assigned a 0.85 probability to a COVID-19 vaccine with greater than or equal to 70% efficacy occurring after 2020. The first vaccine to show an efficacy greater than or equal to 70% was the Pfizer-BioNTech vaccine, reporting an efficacy of 95% on Dec. 10, 2020.

The consensus median prediction made in June, 2020 for the efficacy of the ChAdOx1 vaccine is 55% with a probability of 0.27 assigned to values between 60% and 80% and a probability of 0.13 assigned to an efficacy below 10% (Fig. 2B.). The reported efficacy of the ChAdOx1 vaccine was 61.2% [70].

The consensus mode prediction of efficacy made in July, 2020 was 60% for a vaccine produced using a non-replicating viral platform, 65% for a vaccine produced using a DNA/RNA platform, 75% for a inactivated, and 77% for a protein sub-unit platform (Fig. 2C.). The probability assigned to an efficacy below 50% was 0.35 for a non-rep platform, 0.30 for a DNA/RNA platform, 0.25 for an inactivated platform, 0.19 for a protein sub-unit platform.

The consensus mode (median) prediction made in August, 2020 of the efficacy of a vaccine approved under a standard regulatory process was 66% (66%) and was 52% (50%) for a vaccine approved under an emergency use authorization (such as Operation Warp Speed) (Fig. 2D.). An efficacy of less than or equal to 50% was assigned by the consensus a probability of 0.16 for a vaccine approved under a standard regulatory process and a probability of 0.51 for a vaccine approved under an emergency use authorization.

### III.3. Safety

The consensus median prediction, made on June 30th, for when a COVID therapy will show a significant difference in survival was March, 2021 (80CI: [Sept., 2020, Dec., 2022]), on July 27th the consensus mode prediction for when a significant survival effect would be reported for a therapy using monoclonal antibodies was Dec., 2020 (80CI: [Nov., 2020, March, 2023]) compared to a mode of Dec., 2020 (80CI: [Oct., 2020, June, 2022]) for an antiviral platform, and June, 2024 (80CI: [Feb., 2021, March, 2024]) for an orally administered treatment. The consensus assigned a probability of 0.67 to a greater than 80% chance of 10 or more serious adverse events caused by a SARS-CoV-2 vaccine (under a standard or emergency regulatory process). We asked trained forecasters and experts six questions, one in June, two in July, one in August, and two in September related to the safety of a COVID-19 vaccine (Fig. 3) and received 131 predictions (21 predictions per question on average)

**FIG. 3:**
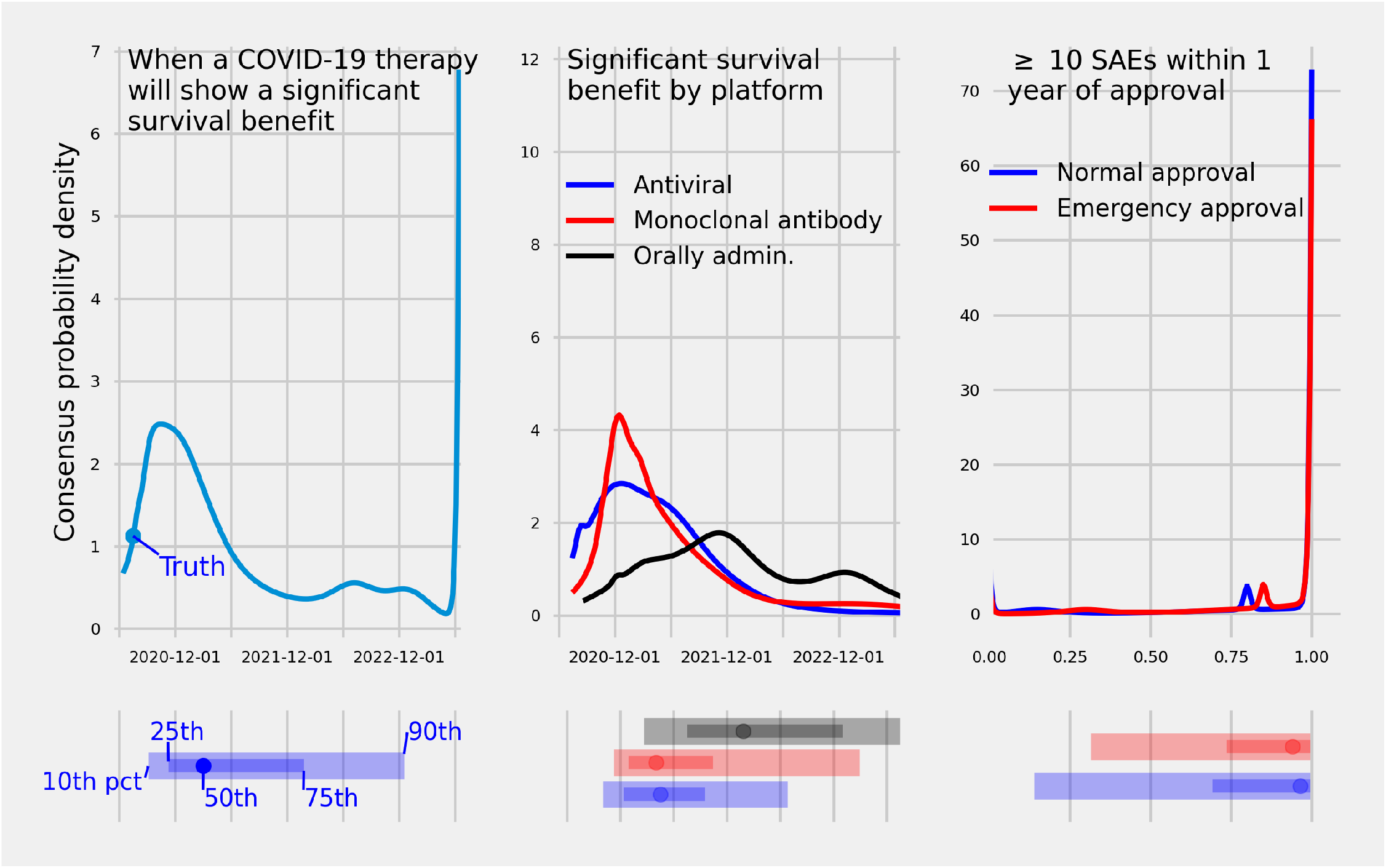
(A.) A consensus predictive density made in June, 2020 over dates for when a COVID-19 therapy will show a significant survival benefit in a randomized clinical trial enrolling more than 200 patients. (B.) Consensus predictive densities made in Aug., 2020 over dates when a SARS-CoV-2 vaccine will show a significant survival benefit in a randomized control trial enrolling more than 200 patients across three different viral platforms. (C.) Consensus predictive densities over the probability of ten or more serious adverse events within one year of the date of approval of the first SARS-CoV-2 vaccine approved through a standard regulatory process or emergency use authorization. Below each density is the 10^th^, 25^th^, 50^th^ (median), 75^th^, and 90^th^ percentile. True values, if available, are represented as a filled circle. The consensus was uncertain when a vaccine would show a survival benefit, assigning 80% confidence intervals that spanned close to two years for when a COVID-19 therapy would show a benefit. The consensus was certain at least 10 SAEs would be observed within one year of approval.

The mode prediction made in June, 2020 of when a COVID-19 therapy would show a significant survival benefit was Oct., 2020 (Fig. 3A.). A probability of 0.35 was assigned to a vaccine showing a survival benefit from June 15th, 2020 to Dec. 31, 2020, a probability of 0.42 was assigned between the dates Jan. 1, 2021 and Dec. 31, 2021, and a probability of 0.23 after 2021. July 17th, 2020 was the 6^th^ percentile of the consensus predictive distribution. Dexamethasone—an anti-inflammatory treatment—was shown to have a significant benefit to survival on July 17th 2020 [71].

The consensus predictive density for when a monoclonal antibody platform would show a significant survival benefit had the smallest interquartile range (IQR: [Dec. 29, 2020, Oct. 15, 2021]), second smallest IQR was for an antiviral platform (IQR: [Dec. 14, 2020, Sept. 16, 2021]), and largest IQR was for an orally administered therapy (IQR: [July, 2021, Jan., 2023]) (Fig 3B.). For the monoclonal antibody and antiviral platform, a 0.28 and 0.30 probability (respectively) was assigned to a survival benefit reported before Jan., 2021. The median prediction for when a monoclonal antibody platform would show a significant survival benefit was April, 2021, for an antiviral platform was April, 2021, and for an orally administered therapy was Jan., 2022.

The consensus median prediction, made on Sept. 30th, 2020, of the probability that greater than or equal to 10 serious adverse events are caused by a vaccine was 0.97 for a vaccine approved under standard regulatory conditions and 0.94 for a vaccine approved under an emergency use authorization.

### III.4. Time to approval

A consensus of experts and trained forecasters made a median prediction in July, 2020 that a SARS-CoV-2 vaccine will be approved in the US or European Union in April, 2021 (80CI: [Oct., 2020, July, 2022]). In August the consensus median prediction for when a vaccine will be approved in the US under an emergency use authorization was Feb., 2021 (80CI: [Sept., 2020, Sept., 2022]) and in September the median prediction was Jan., 2021 (80CI: [Oct., 2020, Nov., 2021]) (Fig. 4). We asked trained forecasters and experts six questions, one in June, one in July, two in August, and two in September related to the timing of approval of a COVID-19 vaccine (Fig. 4) and received 120 predictions (20 predictions per question on average)

**FIG. 4:**
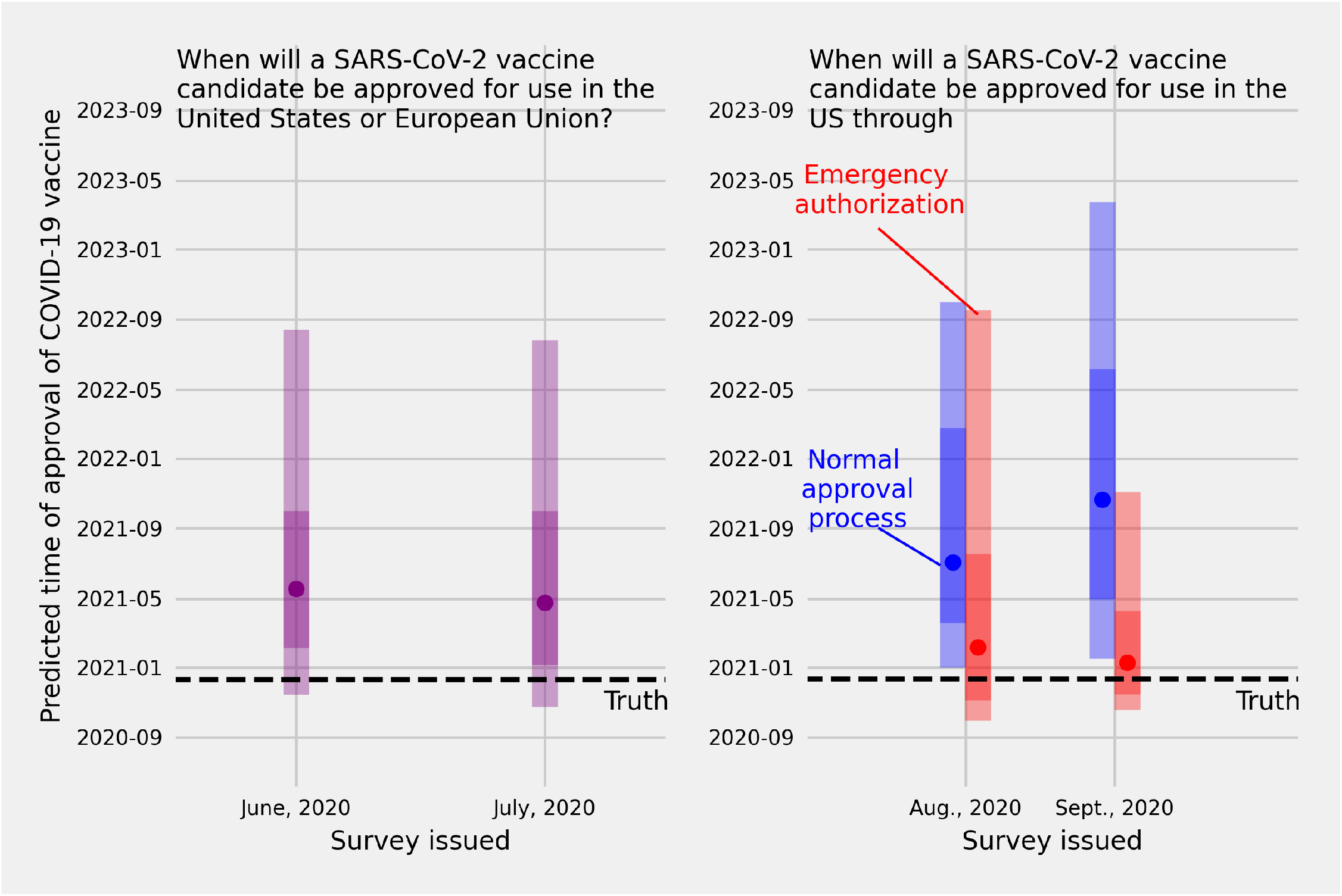
(A.) Consensus predictive percentiles made in June and in July, 2020 for the date when SARS-CoV-2 vaccine will be approved for use in the US or European Union. (B.) Consensus predictive percentiles for the date a SARS-CoV-2 vaccine will be approved for use in the US through a standard regulatory process (blue) or an emergency use authorization (red). The consensus median predictions made in June and July for when a SARS-CoV-2 candidate would be approved in the US or EU were many months later than the truth (May, 2020 and April, 2020 vs Dec., 2020). Consensus median predictions of the date of emergency approval of a SARS-CoV-2 vaccine in the US were Feb., 2020 (made in Aug., 2020) and Jan., 2020 (made in Sept., 2020) which were close to the true date Dec., 2020. An environmental cue, time between when the forecast was made and the truth, or how the question was asked, may have impacted predictive accuracy.

The consensus median prediction made on June 30th, 2020 for when a SARS-CoV-2 vaccine will be approved—either under a standard regulatory process or an emergency use authorization—in either the US or the European Union was May, 2021 (80CI: [Nov., 2020, Aug., 2022]) and the median prediction made on July 31st, 2020 was April, 2021 (80CI: [Oct., 2020, July, 2022]) (Fig. 4A.). There was a small difference in the median prediction between the survey issued in June and the survey issued in July (Diff. = 25 days), and a small difference in the number of days between the lower and upper bound of the 80% confidence interval made in June (637 days) and in July (639 days).

The consensus median prediction made in August, 2020 of when a SARS-CoV-2 vaccine would be approved in the US and under an emergency use authorization was Feb., 2021 (80CI: [Sept., 2020, Sept., 2022]) and for the consensus prediction made in September, 2020 was Jan., 2021 (80CI: [Oct., 2020, Nov., 2021]) (Fig. 4B. red). Under a standard regulatory process the consensus median prediction made in August, 2020 of when a SARS-CoV-2 vaccine would be approved was July, 2021 (80CI: [Dec.,2020, Oct., 2022]) and the consensus prediction made in September, 2020 was Oct., 2021 (80CI: [Jan., 2021, March, 2023]) (Fig. 4B. blue)

The difference in the days between the median prediction of when a vaccine candidate would be approved under an emergency use authorization vs a normal regulatory process was 148 days in August and 285 days in September. The September median prediction compared to the August median prediction of when a vaccine would be approved under an emergency use authorization was 26 days sooner (from Feb, 2020 to Jan 2020) and the September median prediction of when a vaccine would be approved under a normal regulatory process was 109 days later (from July to October, 2021).

### III.5. Rate of production and delivery

The consensus median prediction made in July, 2020 of when an approved vaccine in the US or EU would be administered to more than 100,000 people was Sept. 2021 (80CI: [Jan., 2021, Oct., 2022]). In Aug, 2020 the median prediction for the number of weeks after approval to produce 100M doses of a vaccine that uses a DNA/RNA platform was 18 (80CI: [5, 51]) and using a viral vector platform was 34 (80CI: [10, 72]) (Fig. 5). We asked trained forecasters and experts three questions, one in June and two in August related to speed to produce and administer a vaccine after (Fig. 5), and received 53 predictions (17 predictions per question on average)

**FIG. 5:**
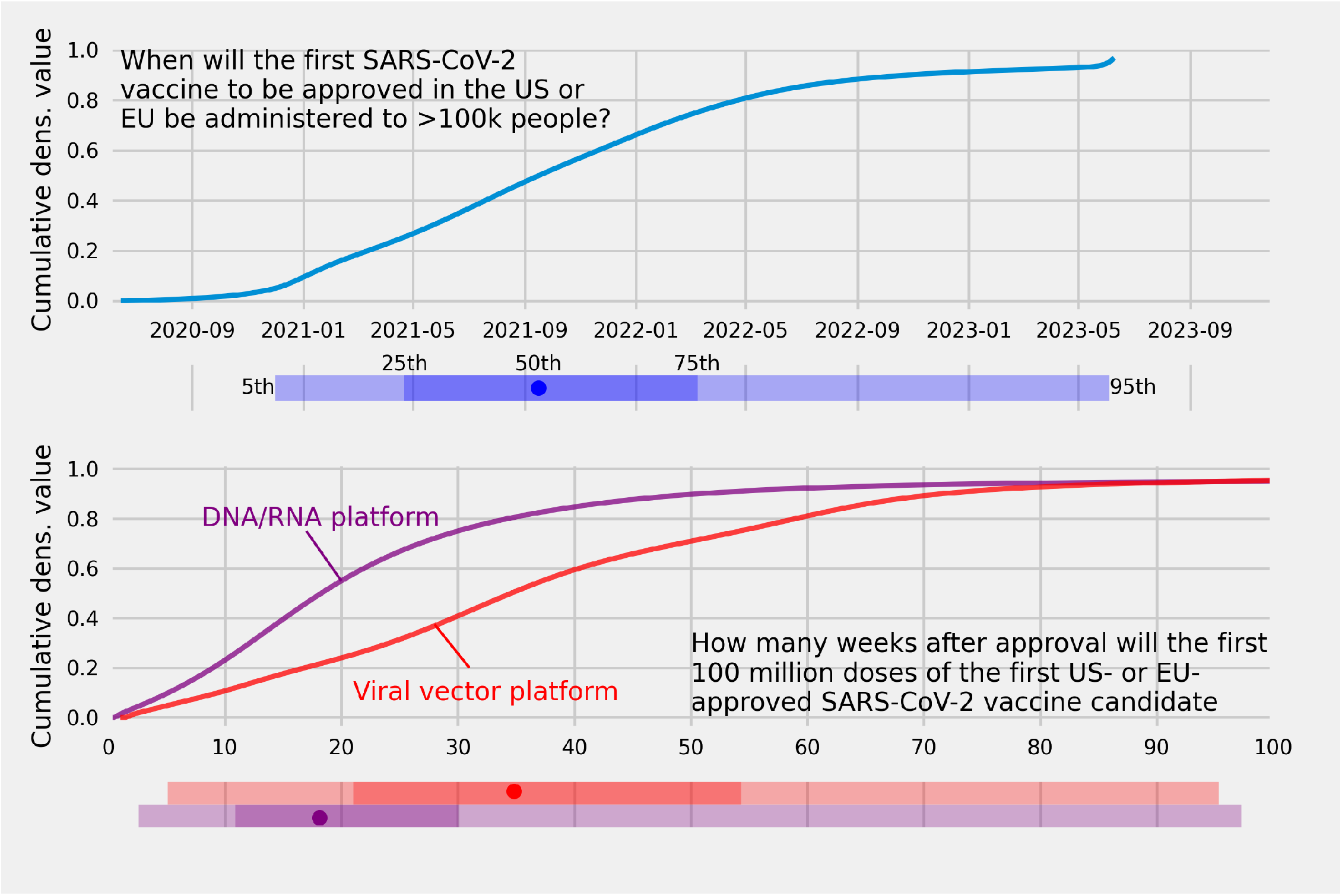
(A.) A consensus cumulative predictive density of trained forecasters and experts over dates for when an approved SARS-CoV-2 vaccine in the US or European Union will be administered to more than 100,000 people. (B.) Consensus cumulative predictive densities of the number of weeks after approval needed to manufacture 100,000,000 doses of a vaccine using a DNA/RNA platform (purple dashed line) and a vaccine using a viral vector platform (red solid line).

The consensus assigned a 0.10 probability to 100K doses of a vaccine administered by the end of 2020 and a 0.66 probability to 100K doses administered by the end of 2021 (Fig. 5A.).

The consensus assigned a 0.27 probability to 10 weeks to manufacture 100M doses of a vaccine using a DNA/RNA platform compared to a 0.10 probability assigned to 10 weeks to produce 100M doses of a vaccine using a viral platform (Fig. 5B.). The difference in the median consensus prediction for the number of weeks to produce 100M doses for a vaccine using a DNA/RNA platform compared to a viral vector platform was 16 weeks.

### III.6. Predictive accuracy of individuals and the consensus

The accuracy of a consensus of trained forecasters plus experts was in between the accuracy of a consensus generated from trained forecasters and the accuracy of a consensus generated from experts except for a single question where individual experts’ accuracy was either below the 25th percentile or above the 75th percentile (Fig. 6). Across all questions where the truth could be determined, the scaled ranks for a consensus of trained forecasters plus experts had a smaller interquartile range when compared to individual forecasters (Fig. 7).

**FIG. 6:**
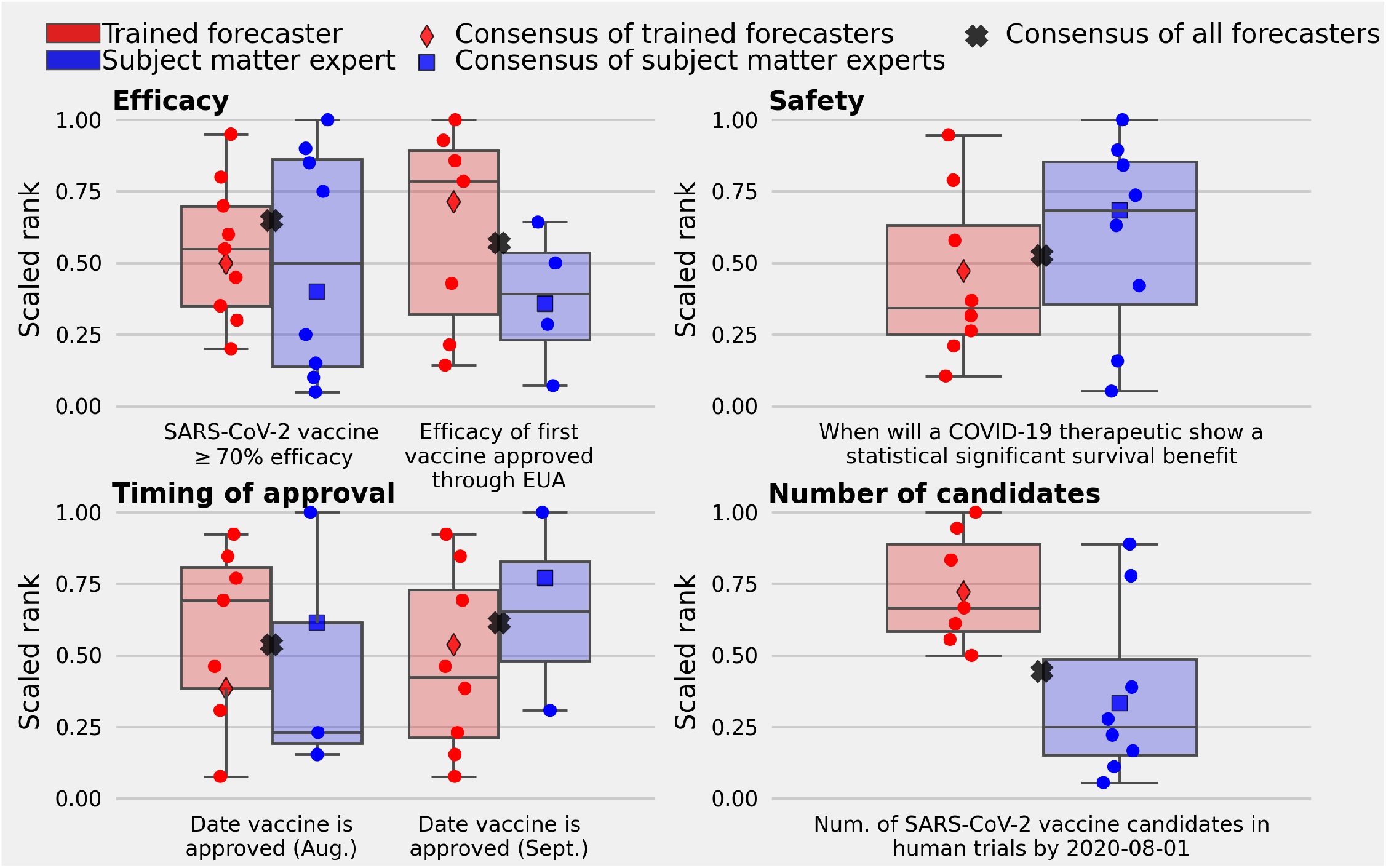
The scaled rank for individual trained forecasters (red circles), subject matter experts (blue circles), and three consensus distributions: (i) a consensus of trained forecasters (red diamond), of subject matter experts (blue square), and the consensus of both trained forecasters and experts (black X) for 6 questions with ground truth. (A.) Two questions with ground truth relate to efficacy, (B.) one question is related to safety, (C.) two questions are related to the timing of vaccine approval, and (D.) one question asked forecasters for the number of vaccine candidates in human trials by Aug., 2020. The mean scaled rank of a consensus of trained forecasters and subject matter experts is 0.58 (80CI: [0.49, 0.63]), a consensus of trained forecasters only is 0.56 (80CI: [0.43, 0.72]), and a consensus of experts only is 0.53 (80CI: [0.35, 0.73]). None of the three consensus predictions are the most accurate, however the trained forecaster plus expert consensus is in the top 50^th^ percentile for all but one question with ground truth. Aggregating trained forecasters and subject matter experts has the potential to guard against an individual forecast with poor accuracy.

**FIG. 7:**
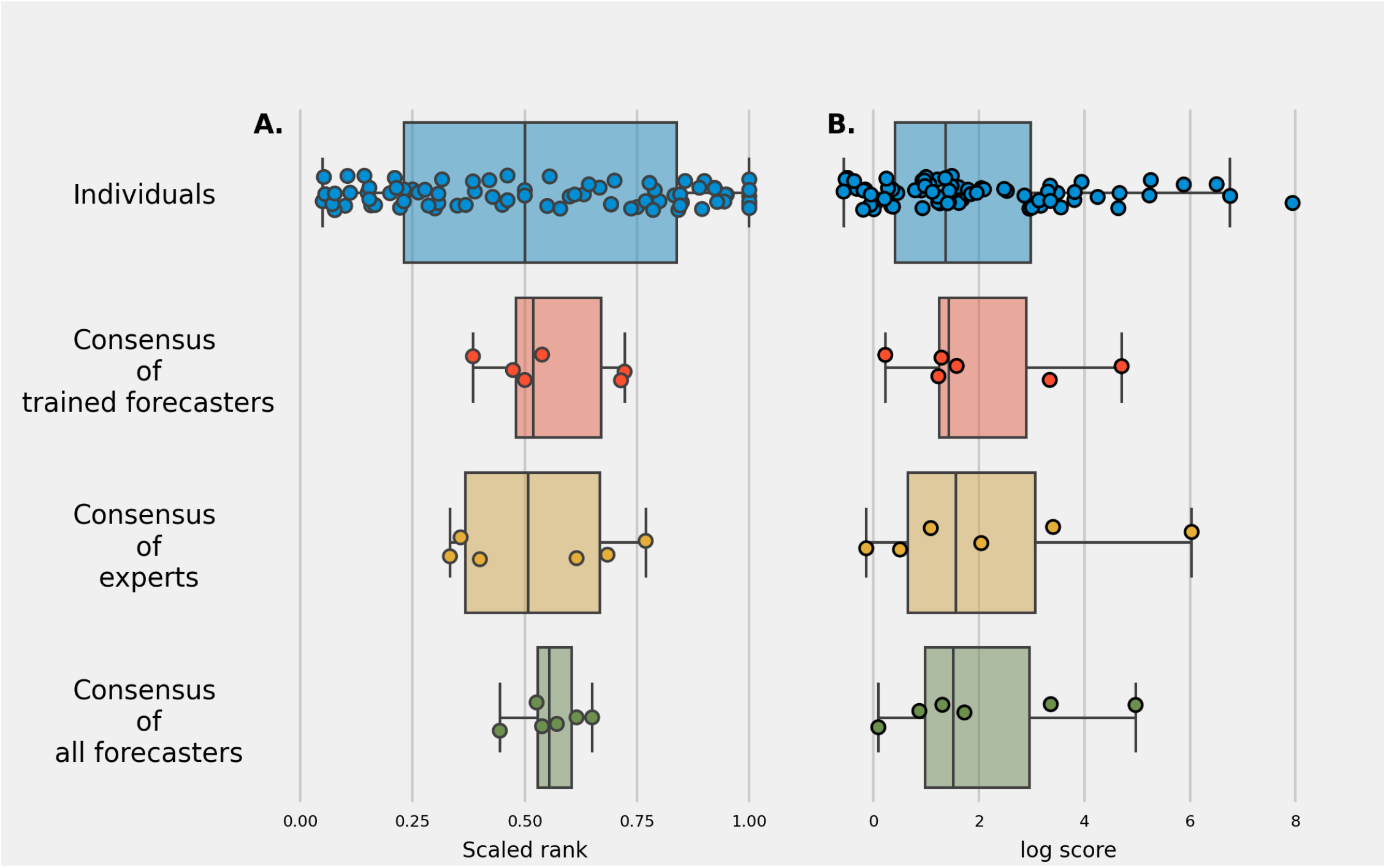
(A.) The scaled rank across all questions where the truth could be determined for individual trained forecasters and experts, a consensus of trained forecasters, consensus of experts, and consensus of both trained forecasters and experts. (B.) The log score across all questions for individuals and the same three consensus predictions. Individual ranks and scores have a larger variability than any of the three consensus forecasts. The consensus of trained forecasters and experts has the smallest interquartile range for scaled rank and second smallest interquartile range for log score. A consensus forecast is often in the top 50th most accurate forecasts and has consistent performance.

The mean scaled rank for individual trained forecasters was 0.56 (80CI: [0.18, 0.94]) and for individual subject matter experts was 0.48 (80CI: [0.08, 0.98]). The standard deviation of the scaled rank was 0.29 for trained forecasters and 0.34 for experts.

The mean scaled rank for the consensus generated from both trained forecasters and experts was 0.58 (80CI: [0.49, 0.63]) compared to a mean scaled rank of a trained forecasters only consensus of 0.56 (80CI: [0.43, 0.72]), and for an experts only consensus of 0.53 (80CI: [0.35, 0.73]). The consensus of trained forecasters and experts was more accurate than both the consensus of trained forecasters and experts for 1/6 questions (proportion: 0.16, 80CI: [0, 0.33]). Compared to a consensus of trained forecasters, a consensus of experts was more accurate for 3/6 questions (proportion: 0.50, 80CI: [0.16, 0.83]). A trained forecaster consensus was more accurate than a consensus of experts for 2/2 efficacy questions (Fig. 6A.) and was more accurate when asked to predict the number of SARS-CoV-2 vaccine candidates that will be in human trials by Aug 1st, 2020 (Fig. 6D.). An expert consensus was more accurate than a consensus of trained forecasters when asked for the date when a COVID-19 therapy will show a survival benefit (Fig. 6B.) and for 2/2 questions related to the timing of approval of a SARS-CoV-2 vaccine.

The 25^th^ and 75^th^ percentiles for scaled rank was, from the widest interval to the smallest interval, [0.23, 0.84] for individuals, [0.48, 0.67] for a consensus of trained forecasters, [0.37, 0.67] for a consensus of experts, and [0.53, 0.60] for a consensus of trained forecasters plus experts (Fig. 7A.). The 25^th^ and 75^th^ percentiles for log scores, from the largest interval to smallest interval, was [0.42, 2.98] for all individuals, [0.65, 3.07] for a consensus of experts, [0.98, 2.96] for a consensus of trained forecasters plus experts, and [1.24, 2.90] for a consensus of trained forecasters (Fig. 7B.).

A consensus of trained forecasters plus subject matter experts scored above the 50^th^ percentile for 5/6 questions while a trained forecaster consensus and an expert only consensus scored above the 50^th^ percentile on 3/6 questions with ground truth.

Trained forecasters had the highest logscores on average, followed by consensus models, and then subject matter experts (Table I). Ninety five percent confidence intervals around the difference in logscores between subject matter experts and trained forecasters, and between consensus models and trained forecasters were large. We do not have enough data on forecast accuracy to conclude statistical significance at a type I error of 5% that trained forecasters made more accurate predictions than subject matter experts or consensus models.

**TABLE I:** Table of coefficients, 95% confidence intervals, and pvalues for a mixed effects model with log score as the dependent variable, three fixed categorical factors to identify log scores from trained forecasters, subject matter experts, or consensus models, and a random intercept by question. Trained forecasters have the highest average logscore followed by consensus models and subject matter experts. However, there is not enough evidence to conclude that these differences are statistically significant

| Variable | Coefficient | 95CI | pvalue |
| --- | --- | --- | --- |
| Intercept (trained forecasters) | 2.10 | (0.60, 3.60) | 0.006 |
| Consensus models. | -0.01 | (-0.51, 0.50) | 0.983 |
| Subject matter experts | -0.14 | (-0.56, 0.29) | 0.531 |
| $\sigma_{\text{question}}$ | 3.42 | | |

## IV. DISCUSSION

A consensus of trained forecasters and experts in infectious disease and vaccinology provided predictions (i) that quantified uncertainty about the efficacy, safety, and timing of approval for a COVID-19 vaccine from July to September 2020, a time when the vaccine landscape and political atmosphere was rapidly changing; (ii) that were made when data on previous pandemics was sparse, likely relying instead on a combination of objective and subjective information, aspects of past pandemics that may have resembled the COVID-19 pandemic, and intuition; and (iii) that were fast and to the best of our knowledge before other human judgment or computational efforts to predict COVID-19 vaccine characteristics.

Expert and trained forecaster probabilistic predictions underestimated the speed of approval and high efficacy of a SARS-CoV-2 vaccine, and the consensus prediction of the date a COVID-19 treatment would show a survival benefit was much later compared to the truth.

The consensus assigned high probabilities to efficacy values above 70% for the Oxford/AstraZeneca vaccine and to vaccines based on four different vaccine technology platforms [72]. That said, the consensus median prediction made in Aug. 2020 of the efficacy of a vaccine approved under emergency authorization was 50%, greatly underestimating the 95% efficacy of the Pfizer/BioNTech vaccine [29]. In addition to underestimating the efficacy of the first approved vaccine, consensus predictions for all questions related to efficacy assigned positive probabilities to values below 50%, the threshold for approval stated by the FDA.

Consensus forecasts underestimated the speed of vaccine development. The Pfizer/BioNTech vaccine was approved Dec. 10th and on July 17th 2020 Dexamethasone—an anti-inflammatory treatment—was shown to have a significant benefit to survival [71]. The consensus in July, 2020 assigned a small probability to a vaccine approved before or on Dec. 10th, 2020, and the consensus median prediction made in June, 2020 for when a COVID-19 therapy would show a survival benefit was approximately eight months later than the true date. Late predictions of approval and when a vaccine would show a survival benefit are closer to the timeline of a traditional vaccine and could be because sparse objective data and subjective data caused forecasters to rely on past documentation related to vaccines that went through a standard regulatory pathway.

Often overlooked advantages to human judgment forecasting, compared to computational models, are the ability of a forecaster to transform textual data from the environment into a prediction and output textual data to backup their predictions while also directing other forecasters to potentially important information. For example, forecasters suggested political considerations may influence the timing of the vaccine authorization/approval process in the U.S. with one forecaster pointing out on June 16th that “pressure on the FDA to approve something will be enormous, and will start building once there are even preliminary phase 3 results.” After our August survey closed and before the September survey opened, the President announced a vaccine might be approved before the 2020 election. We feel political events like this could have influenced forecasters because although the median prediction for when a SARS-CoV-2 vaccine will be approved in the US under emergency authorization shifted from Feb, 2021 to Jan, 2021 (One month sooner) in the midst of broadcasted political pressure about approving a vaccine, the median prediction for a vaccine to be approved under a standard regulatory process moved from July, 2021 to Oct., 2021 (3 months later). Humans are better positioned than computational models to synthesize textual data into their forecasts and, unlike computational models, can generate commentary that may present new information to others who plan to generate a forecast. Though the ability of forecasters to process textual information is an advantage in many cases, it can also mislead forecasts.

Individual predictive accuracy was higher for trained forecasters than for subject matter experts, and we found that aggregating individual predictions into a consensus can guard against inaccurate predictions made by individuals. Forecaster’s individual accuracy was inconsistent and ranged from the bottom 25^th^ percentile to the top 75th percentile across question type (efficacy, safety, timing) and from survey to survey. Compared to subject matter experts, trained forecasters had on average higher logscores. This may suggest predictive accuracy may depend more on forming good forecasting habits than on the subject matter.

Though a consensus was never the most accurate prediction, a consensus of trained forecasters and experts was in the top 50th percentile for five out of six questions. A consensus may not be the most accurate forecast, but a consensus can potentially guard against individual forecasters with poor accuracy.

In this work we are limited by a small number of questions we could ask forecasters, the number of questions that could be compared to the truth, and difficulties associated with human judgment.

A computational model, if possible to build, would be likely be able to make predictions for a large number of similar questions. However a forecaster often spends a significant amount of time to generate a forecast.

Only 23 percent (6/26) of the questions resolved, having true values we can use to to assess forecaster accuracy. A subset of questions will resolve over time but many questions will never resolve because the criteria to determine the truth (to resolve the question) may have been too strict. Many questions asked forecasters to predict the efficacy, safety, and timing of a vaccine that is granted a biologics license application (BLA), however, to date no SARS-CoV-2 vaccine has been granted a BLA.

The number of questions that are unable to resolve highlights that those who pose the questions are as important as those who submits predictions. Before forecasters begin to submit predictions, the author, when developing the question, must first predict whether or not they expect to obtain ground truth. For instance, there was uncertainty about whether or not the standard authorization process, emergency authorization process, or both would be used to approve vaccines. We expected vaccines to be approved by both pathways, but because only the emergency authorization process has been used any question limited to the standard regulatory process will never have a ground truth.

Forecasters were able to submit comments along with quantitative predictions, but a small percentage (12%, 63 comments divided by 530 predictions) of forecasters accompanied their predictions with text that explained the data they relied on and the rationale/model used to generate a probabilistic prediction. An ongoing challenge in human judgment forecasting is to solicit a prediction, data used to make the prediction, and a rationale.

Forecasts were made by trained forecasters and subject matter experts in a rapidly changing, chaotic environment and so, in line with representative design, we suggest our results can only be generalized to human judgment forecasting in a noisy environment without a large volume of historical data related to the questions asked of forecasters [73].

The purpose of this work was to develop predictive guidance for the public and public health officials and not to rigorously evaluate forecaster accuracy. Further experimental work is needed comparing subject matter experts to trained forecasters while varying task predictability and the number of linear and non-linear cues available in the environment to the forecaster. This work weighted equally predictive distributions from subject matter experts and trained forecasters, and further work is needed that explores differential weighting algorithms that may produce better accuracy.

Human judgement forecasting of progress towards a vaccine can be viewed as a tool to support primary preventative measures against an infectious agent. Probabilistic forecasts can target multiple audiences to support the public when making complex decisions about their health under uncertainty and support both short term and long-term health promotion practices implemented by public health officials. Because vaccination is a low frequency, voluntary event, forecasts did not aim to modify the public’s sense of control or desire to engage in inoculation. Instead, forecasts made easily accessible online aimed to help ready individuals at all risk levels for vaccination and understand the impact of their decisions on their own and on other’s health. Predictions from a consensus of the efficacy, safety, and timing of a vaccine may have been accurate enough to improve situational awareness for public health officials.

Consensus predictions based on human judgment are a promising tool to aid decision-making under the uncertainty of a rapidly evolving situation where there is little historical data, where data is only loosely applicable, and when timely risk communication is important.

## Data Availability

As much data and code as possible is available at https://github.com/computationalUncertaintyLab/vaccinceAndTherapeuticsCrowd

## VII. SUPPLEMENTAL MATERIALS

### VIII. QUESTIONS ASKED FOR ALL FOUR SURVEYS

**FIG. S1:**
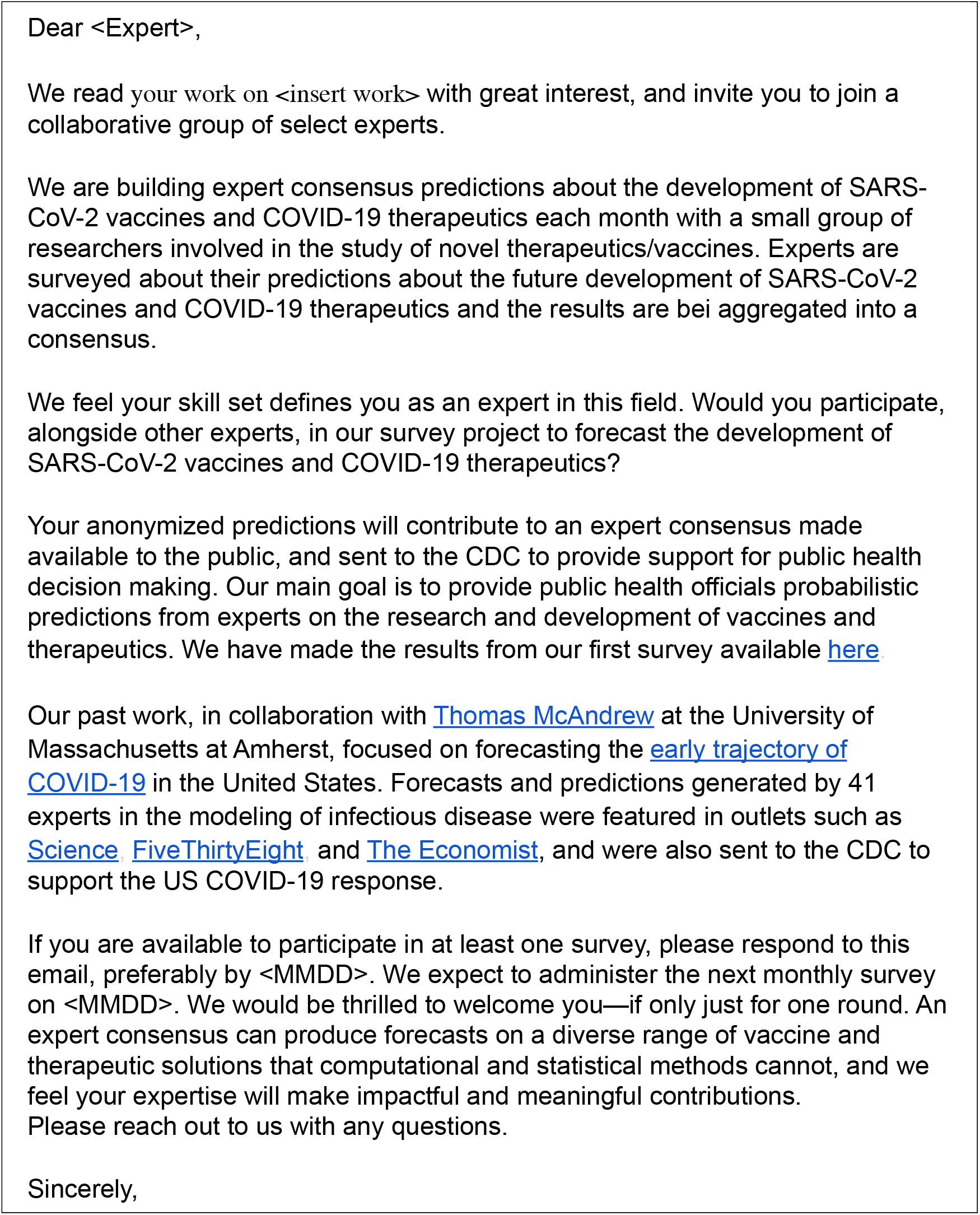
A template email used to solicit forecasting participation from subject matter experts in molecular and cellular biology, microbiology, virology, biochemistry, and infectious disease who have had several years of experience studying vaccine, antiviral, or biological related to infectious agents.

**TABLE S1:**
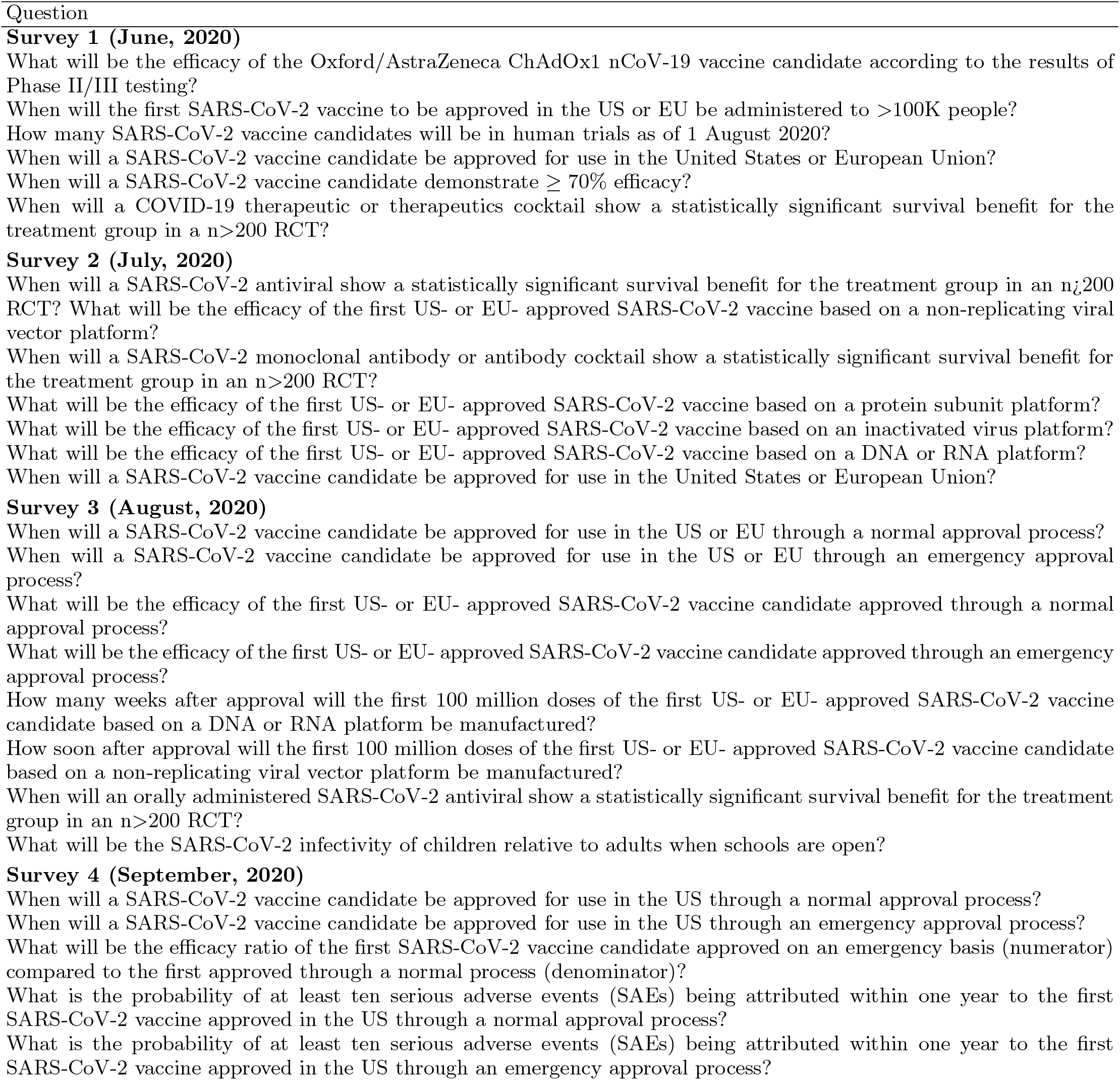
List of all questions stratified by survey.

